# Causal effects from non-alcoholic fatty liver disease on kidney function: A Mendelian randomization study

**DOI:** 10.1101/2021.02.22.21252263

**Authors:** Sehoon Park, Soojin Lee, Yaerim Kim, Semin Cho, Kwangsoo Kim, Yong Chul Kim, Seung Seok Han, Hajeong Lee, Jung Pyo Lee, Kwon Wook Joo, Chun Soo Lim, Yon Su Kim, Dong Ki Kim

## Abstract

**Background & aims:** An observational association between nonalcoholic fatty liver disease (NAFLD) and kidney function impairment has been reported. A genetic variant linked to an increased risk of NAFLD, the G allele of rs738409, has been reported to be associated with a reduction in estimated glomerular filtration rate (eGFR).

**Approach & Results:** In this Mendelian randomization (MR) study, we first performed single-variant MR with rs738409 as a genetic instrument to predict NAFLD. Another genetic instrument was developed from a previous genome-wide association study for the NAFLD phenotype in the Million Veteran Program cohort among individuals of European ancestry (68,725 cases and 95,472 controls). The eGFR outcome was assessed in individuals of white British ancestry included in the UK Biobank (N = 321,405). Further, the associations were reassessed in two negative control subgroups (body mass index < 25 kg/m^2^ and serum alanine aminotransferase level < 20 IU/mL) with a low probability of developing NAFLD. As a replication analysis, a summary-level MR was performed with the European ancestry CKDGen dataset (N = 567,460). In the UK Biobank dataset, a genetic predisposition for NAFLD, either by rs738409 or a group of variants, was significantly associated with a reduced eGFR even with adjustment for major metabolic disorders. Although the associations were not significant in the negative control subgroups with a low probability of developing NAFLD, they were significant in the subgroups with a remaining risk of NAFLD, suggesting the absence of a horizontal pleiotropic pathway. The summary-level MR from the CKDGen dataset supported the causal effects of NAFLD on reduced eGFR.

**Conclusions:** This MR analysis supports the causal reduction in kidney function by NAFLD.

---

Nonalcoholic fatty liver disease (NAFLD) is one of the leading comorbidities worldwide and places a large socioeconomic burden on the population.^1^ The prevalence of NAFLD has increased as the population with obesity has grown. Obesity and related metabolic disorders contribute to NAFLD development, and genetic factors also play roles in NAFLD and its severity.

Chronic kidney disease (CKD) is another major comorbidity with a large socioeconomic burden, affecting nearly 7 million individuals in the global population.^2^ Previous studies reported that NAFLD may cause kidney function impairment, which is supported by a recent systematic meta-analysis of observational findings.^3,4^ However, as the risks of NAFLD and CKD are both affected by obesity-related metabolic disorders, whether NAFLD has a direct causal effect on kidney function cannot be inferred from conventional observational findings that are inevitably affected by unmeasured confounding effects or reverse causation. Therefore, additional investigation that may evaluate the causal effects of NAFLD on kidney function is warranted. Such evidence would suggest the need to monitor kidney function in NAFLD patients and the possibility that appropriate management of NAFLD may lead to the prevention of kidney function impairment.

Previous studies identified that the PNPLA3 gene single nucleotide polymorphism (SNP), rs738409, is a genetic causal variant for NAFLD with the strongest association strength.^5^ The G allele of rs738409 has been reported to be associated with NAFLD in genome-wide association studies (GWASs) and to cause cirrhotic changes in the liver in NAFLD patients. Interestingly, previous reports identified that rs738409 is also associated with kidney function in several cohorts and by experimental findings.^6-10^ This may suggest a causal association between the two disorders; however, a further large-scale study is warranted to dissect this association between rs738409 and NAFLD or kidney function to demonstrate the causal direction.

Mendelian randomization (MR) is an analytic tool for estimating the causal effect of an exposure on complex outcomes.^11^ As genotype is fixed before birth, genetically predicted exposure is minimally affected by reverse causation or confounding effects. The method has been implemented in the recent medical literature and identified important causal biological pathways between complex disorders. In addition, with the availability of large-scale genetic data for kidney function traits, MR has been utilized to identify causal risk factors for kidney function impairment.^12-15^

In this MR study, we first performed MR analysis by selecting rs738409 as a genetic instrument to predict NAFLD and assess its association with kidney function outcome in two independent, population-scale genetic datasets for kidney function traits. We also introduced an alternate genetic instrument including a group of SNPs to apply recently popularized MR methodology in our analysis. We hypothesized that causal effects of NAFLD on kidney function parameter, estimated glomerular filtration rate (eGFR), would be present.

## Materials and Methods

### Ethical considerations

This study was approved by the Institutional Review Boards of Seoul National University Hospital (E-2010-007-1160). The requirement for informed consent was waived as we investigated anonymous public databases. The investigation of the UK Biobank was approved by the consortium (No. 53799).

### Concept of MR analysis

Unlike direct assessment of the effect of a SNP, MR tests the association between genetically predicted exposure and an outcome, which would be mediated by phenotypical exposure (Figure 1).^11^ As not all individuals with a genetic predisposition develop the phenotype of interest, MR requires a large study sample to demonstrate causal estimates. The randomization process occurs before birth, and MR stratifies the population according to their genetic risk for exposure. For instance, as the rs738409 G allele has been identified to be the most significant genetic variant for NAFLD, those with the G/G rs738409 genotype would have a higher probability of developing NAFLD than those with the C/C genotype. Thus, if individuals with the rs738409 G allele have worse kidney function than those carrying other alleles, the genetic effect of the G allele on NAFLD would suggest a causal effect of NAFLD on kidney function. As one’s genotype precedes the development of clinical confounders or reverse causation, a causal inference of the identified association is possible. To validly demonstrate a causal effect, MR requires attaining three key assumptions: the relevance assumption, independence assumption, and exclusion-restriction assumption. The relevance assumption is that the genetic instrument should be strongly associated with the exposure of interest. The independence assumption and exclusion-restriction assumption indicate the absence of a horizontal pleiotropic pathway, and the assumptions are violated when the genetic effect from an instrument is not through the exposure of interest but through a confounder or another mechanism.

**Figure 1.**
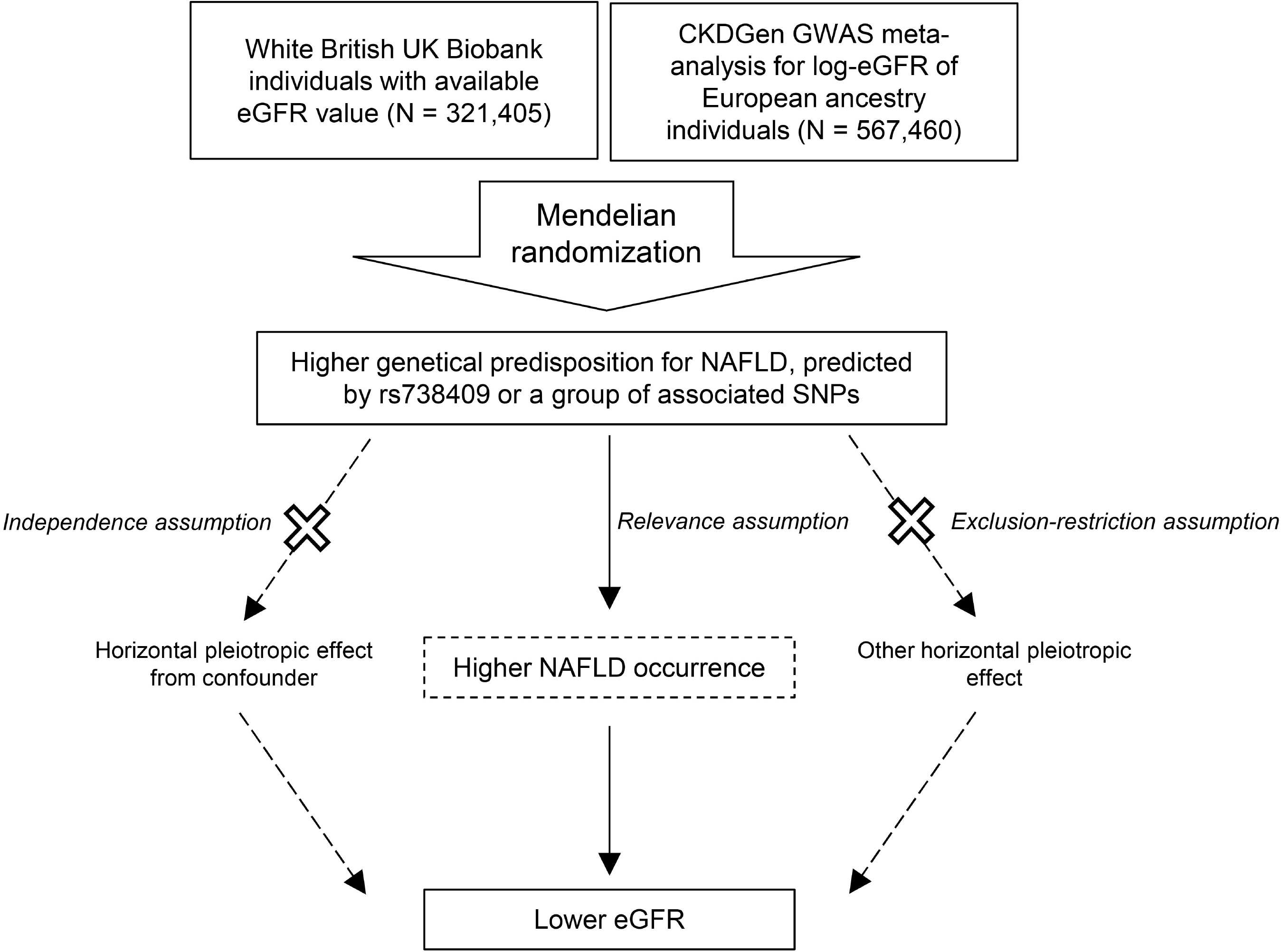
Study flow diagram. The study included individual-level Mendelian randomization analysis with the UK Biobank and summary-levle Mendelian randomization analysis with the CKDGen data. To deomnstrate causal estiamtes from genetically predicted exposures, Mendelian randomization analysis requires to attain three key assumptions. The relevance assumption is that the genetic instrument should be strongly associated with the exposure of interest, and this assumption was considered attained as the study instrumented two most significant SNPs from previous genome-wide assiciation studies. The independence assumption is that the genetic effect should not be from a confounder. The exclusion-restriction assumptions means that there is no horizontal pleiotropic pathway other than a confounder (e.g. direct effect from a SNP to outcome).

### Genetic instruments for NAFLD

The abovementioned SNP rs738409 is a confirmed causal variant for NAFLD and is also associated with disease severity, including steatosis, fibrosis, and NAFLD activity score.^5^ As the SNP genetically explained NAFLD to the largest degree with confirmed direct causality, we first performed a single-variant MR with the SNP. Although including a large number of variants to genetically predict a phenotype in MR has value for increasing the explained variance by genotype, the issue of horizontal pleiotropy increases according to the method.

This issue was of particular concern because other SNPs reported in a previous GWAS were associated with metabolism-related phenotypes rather than with NAFLD.^5^ Considering that kidney function and NAFLD are both closely related to metabolism-related phenotypes, we first performed MR analysis with single-variant MR, which has the advantage of demonstrating specific causal effects and minimizing the effects of the horizontal pleiotropic pathway. This was also because of the particular interest in rs738409, considering previous reports for its’ association with kidney function.

Next, to include other relevant variants for NAFLD and to implement an analysis with a recently popularized method involving pleiotropy-robust MR sensitivity analysis of a group of instrumented SNPs, which has strength reducing the possibility of weak-instrument bias, we used the results from the current largest-scale GWAS for clinically diagnosed NAFLD in the Million Veteran Program by Vojkovic M et al.^16,17^ We also considered another GWAS with histologically confirmed NAFLD, but the reported 6 genetic loci were mostly associated with obesity or metabolism-related traits, raising concerns of a pleiotropic effect; thus, we implemented the larger-scale GWAS with the Million Veteran Program cohort.^5,17^ To minimize the effects of different ethnic distributions, we screened 55 SNPs from the European ancestry-specific analysis, including 68,725 NAFLD cases and 95,472 controls (Supplemental Table 1). From Steiger filtering, we identified a SNP (rs12500824) with a reverse-direction effect (association with eGFR outcome, P = 2.61E-61 and with NAFLD exposure, P = 1.37E-9);^18^ thus, the SNP was disregarded. Regarding a horizontal pleiotropic effect from shared metabolic/circulatory risk factors between kidney function and NAFLD, we disregarded 14 SNPs that had a strong association with any phenotype of the circulatory system (e.g., hypertension) or endocrine/metabolism category (e.g., body mass index or dyslipidemia) in the phenome-wide association study with the UK Biobank data reported in a previous study (Supplemental Table 2).^5^ Thus, 40 SNPs remained as the genetic instrument with a group of SNPs for NAFLD. The genetic instrument developed from the Million Veteran Program data had particular strength, as the cohort was independent from the outcome data (UK Biobank and CKDGen) in this study, enabling us to perform two-sample MR analysis, which has strength due to its low possibility of false-positive findings.

### Considerations for MR assumptions

The relevance assumption was considered attained, as the single variant MR implemented the genetic variants with strongest association with NAFLD or liver steatohepatitis. Also, the alternate genetic instrument for NAFLD consisting a group of variants was also reported from a large-scale GWAS study with association reaching genome-wide significant level.

The independence assumption was considered attained, particularly in the single-SNP MR analysis because considering the direct association with the variants with NAFLD or related phenotypes, the effect from the SNP would minimally from a confounding factor. In the genetic instrument including multiple SNPs, we carefully considered the confounding effects from metabolic disease-related phenotypes and disregarded the variants related to such confounders as those described above. In addition, we performed several pleiotropy-robust MR sensitivity analyses to support the attainment of the independence assumption.

Along with the consideration for exclusion-restriction assumption, in the current MR analysis, we paid additional attention to the possibility of a horizontal pleiotropic pathway, as some reports, despite the inclusion of only a few hundred individuals, suggest that rs738409 may directly affect kidney function.^6,8,9^ If the effect from rs738409 on kidney function is direct, than as the genetic effect is not through NAFLD, the causal effect from genetically predicted NAFLD by rs738409 on kidney function could not be studied by MR. Currently, there is no direct statistical test assessing the exclusion-restriction assumption. Therefore, ws suggested in the literature, to test whether there is an independent effect of rs738409 on kidney function, we implemented the “negative control approach”, which has been suggested to be useful for testing the absence of a horizontal pleiotropic pathway when individual-level data is available.^11,19^ The negative control approach re-evaluates the association between genetic instruments and outcome in those with a lower probability of developing the exposure phenotype. For instance, if an estimate of the causal effect of genetically predicted alcohol drinking behavior on a health outcome is significant in a female population, this would be less likely to indicate a true causal effect than the presence of a horizontal pleiotropic pathway. To perform this analysis, we constructed two negative control subgroups with a low probability of having the NAFLD phenotype with the available individual-level data. First, we divided subgroups according to whether the participants had a normal range body mass index (< 25 kg/m^2^), which is not in a range or overweight or obese and would have a small probability of having NAFLD. Second, we divided subgroups according to an alanine aminotransferase level < 20 IU/mL. Although the absolute diagnostic criterion with alanine aminotransferase level has not been determined, it would be reasonable that individuals with a low level would less like to have NAFLD. The association between genetical predisposition for NAFLD and eGFR was reassessed in the negative control group, with minimal risk of NAFLD, and their counterparts, with remaining possibility of NAFLD. If a significant association is identified only in the population with a higher probability of NAFLD, this would support that phenotypical NAFLD is the driving factor of the genetic effects of the instruments on kidney function. Conversely, if the association is similarly identified or more prominent in the negative controls, this would discourage the interpretation that phenotypical NAFLD is the mediating factor from the genetically proxied NAFLD on kidney function. Lastly, the median-based MR sensitivity analysis, which was performed in our summary-level MR analysis with multiple SNPs, relax the assumption in up to half of the genetic variants, thus has been considered as a valid sensitivity analysis for this assumption.

### UK Biobank data source

The UK Biobank is a prospective cohort of > 500,000 individuals aged 50-65 years in the United Kingdom.^20,21^ The database includes invaluable resources of various phenotypical data, and with deeply genotyped information, the UK Biobank data have been implemented in many genetic studies in the current literature. Regarding this study, the database has a particular strength, as other phenotypical data are available, and direct adjustment for kidney function-related metabolic comorbidities was possible. Moreover, the individual-level data enabled us to perform a negative control analysis, which was crucial for supporting the absence of a horizontal pleiotropic pathway.

### MR analysis with the individual-level UK Biobank data

In the analysis, we included 337,138 white British UK Biobank participants passing the sample quality control.^12^ We calculated the eGFR based on cystatin C levels, which has some superiority over creatinine-based values as they are less affected by body shape or diet,^22^ and the values were the outcome variable in the individual-level MR analysis. There were 321,405 individuals with available cystatin C-based eGFR levels. The first exposure was an addition of the G allele in rs738409 as a single-variant MR. The second exposure was the polygenic core (PGS) for NAFLD, calculated from the genetic instrument including 40 NAFLD-associated SNPs by PLINK 2.0. We constructed a regression model including age, sex, body mass index, waist circumference, and the first 10 genetic principal components. In addition, a covariate-adjusted model was constructed and also included diabetes mellitus, medication history for hypertension, systolic blood pressure, diastolic blood pressure, medication history for dyslipidemia, serum triglyceride level, low-density lipoprotein level, high-density lipoprotein level, income grade, and smoking history (Supplemental Methods).

The analysis in the negative control group and the counterparts was also performed. A two-sided P value < 0.05 was considered indicative of a statistically significant finding.^12^

### MR analysis with the CKDGen data

To replicate the findings on a large scale, we demonstrated the causal estimates with the largest GWAS meta-analysis for eGFR performed by the CKDGen consortium.^23^ The outcome summary statistics in the database, which are available in the public domain (URL: https://ckdgen.imbi.uni-freiburg.de/), have been utilized for various MR analyses in the current literature regarding kidney function.^12,13^ The CKDGen GWAS summary result is independent from the UK Biobank data; thus, a similar finding in the data would be an independent replication of the main analysis.

To minimize the effects of different ethnic distributions, we limited the analysis to individuals of European ancestry (N = 567,460). The summary statistics provided in the data were for log-transformed eGFR traits, which were rescaled to a percent change unit.

In the single-variant MR, the effect from addition of the minor alleles of rs738409 on eGFR was tested by the Wald ratio method, as the analysis instrumented a single SNP, with the exposure effect size for NAFLD introduced from a previous GWAS.^5^

In the summary-level MR with multiple instrumented SNPs, we first performed the conventional inverse variance weighted method. Next, pleiotropy-robust MR sensitivity analyses were performed. First, MR-Egger regression with bootstrapped standard error, which provides pleiotropy-corrected causal estimates, was performed with MR-Egger intercept to detect the presence of a pleiotropic effect.^24^ Second, median-based methods, including weighted median and simple median methods, were performed, and such methods have strength because they allow the presence of invalid instruments up to 50%.^25^ Third, the contamination-mixture method was implemented, and this method has strength, as it detects groups of SNPs with similar causal estimates and performs robust MR analysis even in the presence of invalid instruments.^26^ Fourth, MR-PRESSO analysis, which detects and corrects the effects from outliers, was performed.^27^ The analysis was performed with the TwoSampleMR and MendelianRandomization packages in R (version 3.6.2).^28^ The negative control approach analysis was not performed with the CKDGen data, as individual-level data to define subgroups were unavailable.

## Results

### Characteristics of the data sources

The included UK Biobank participants had a median age of 58 [interquartile range 50, 63] years old, and 48% of them were male. The prevalence of obesity and diabetes was 24% and 5%, respectively. There were 20% and 17% of individuals taking medications for blood pressure control and dyslipidemia, respectively. While 5% of the individuals had the highest income grade (> 100000 ₤ per month), 21%, 26%, 26%, and 22% of individuals reported being in the 52000 to 1000000 ₤, 31000 to 51999 ₤, 18000 to 30999 ₤, and < 18000 ₤ income categories, respectively.

Individuals of European ancestry in the CKDGen dataset had a median age of 54 years old, and 50% of them were male, with a median eGFR of 91.4 mL/min/1.73 m^2^ and a prevalence of CKD of 9%.

### MR results in the UK Biobank data

The G allele of rs738409 was significantly associated with reduced eGFR values in the UK Biobank dataset (Table 1). In addition, the PGS for NAFLD showed a significant association with a lower eGFR. When the analysis was restricted to those with normal weight (body mass index < 25 kg/m^2^) and those with a decreased probability of NAFLD development, the association was attenuated and became nonsignificant. However, when the analysis was performed including those with body mass index ≥ 25 kg/m^2^, genetically predicted NAFLD was significantly associated with lower eGFR levels. In another negative control group including those with an alanine aminotransferase level < 20 IU/mL, genetically predicted NAFLD was nonsignificantly associated with eGFR. On the other hand, in those with alanine aminotransferase ≥ 20 IU/mL, who would have an increased probability of NAFLD, the G allele of rs738409 or PGS for NAFLD was significantly associated with reduced eGFR values as the main findings.

**Table 1.** Association between G allele of rs738409 or PGS for NAFLD with eGFR in the individual-level UK Biobank data.

| Study group | Genetic instrument | Base model <sup>a</sup> |  | Covariate-adjusted model <sup>b</sup> |  |
| --- | --- | --- | --- | --- | --- |
|  |  | adjusted beta (95% CI) | P | adjusted beta (95% CI) | P |
| Total white British ancestry individuals (N = 321,405) | G allele of rs738409 | -0.123 (-0.21, -0.036) | 0.005 | -0.104 (-0.203, -0.004) | 0.041 |
|  | PGS for NAFLD | -2.277 (-3.482, -1.071) | < 0.001 | -2.031 (-3.415, -0.647) | 0.004 |
| Negative control subgroups |  |  |  |  |  |
| body mass index < 25 kg/m<sup>2</sup> (N = 106,210) | G allele of rs738409 | -0.023 (-0.173, 0.127) | 0.766 | 0.075 (-0.096, 0.245) | 0.391 |
|  | PGS for NAFLD | -1.823 (-3.913, 0.266) | 0.087 | -0.668 (-3.046, 1.71) | 0.582 |
| ALT < 20 IU/mL (N = 157,928) | G allele of rs738409 | -0.008 (-0.135, 0.12) | 0.906 | -0.016 (-0.161, 0.13) | 0.834 |
|  | PGS for NAFLD | -0.717 (-2.476, 1.042) | 0.424 | -0.846 (-2.782, 1.09) | 0.392 |
| Positive control subgroups |  |  |  |  |  |
| body mass index ≥ 25 kg/m<sup>2</sup> (N = 214,173) | G allele of rs738409 | -0.172 (-0.278, -0.066) | 0.001 | -0.185 (-0.307, -0.063) | 0.003 |
|  | PGS for NAFLD | -2.488 (-3.962, -1.014) | 0.001 | -2.612 (-4.31, -0.914) | 0.003 |
| ALT ≥ 20 IU/mL (N = 163,129) | G allele of rs738409 | -0.237 (-0.355, -0.118) | < 0.001 | -0.183 (-0.319, -0.046) | 0.009 |
|  | PGS for NAFLD | -3.949 (-5.608, -2.291) | < 0.001 | -3.506 (-5.416, -1.596) | 0.001 |

PGS = polygenic score, NAFLD = non-alcoholic fatty liver disease, ALT = alanine aminotransferase
The unit of PGS exposure for NAFLD was one exponential increase and the unit of outcome was continuous cystatin C-based eGFR value (mL/min/1.73 m<sup>2</sup>)
<sup>a</sup> The causal estimates were calculated by linear regression, adjusted for age, sex, body mass index, waist circumference, and the first 10 genetic principal components.
<sup>b</sup> The linear regression model additionally included diabetes mellitus, medication history of hypertension, systolic blood pressure, diastolic blood pressure, medication history for dyslipidemia, triglyceride, high-density lipoprotein, low-density lipoprotein levels, smoking history (never, previous, and current), and income grades.

### MR results in the CKDGen data

In the summary-level MR from the independent CKDGen dataset, NAFLD that was genetically predicted by rs738409 was significantly associated with reduced eGFR values (Table 2). When summary-level MR was performed with multiple SNPs to predict NAFLD, all implemented MR methods, namely, the inverse variance weighted, MR-Egger regression, weighted median, simple median, contamination mixture, and MR-PRESSO methods, all provided significant causal estimates, indicating that genetically predicted NAFLD was significantly associated with reduced eGFR values.

**Table 2.**
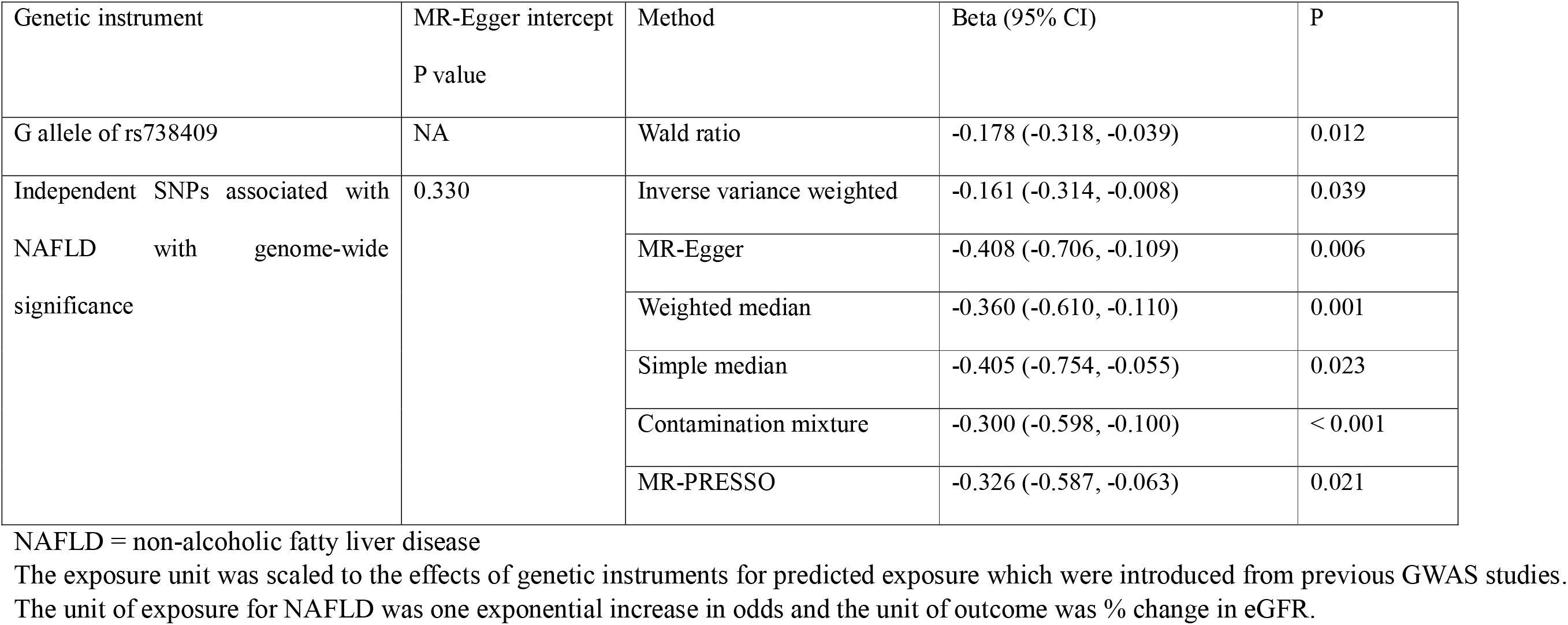
Summary-level Mendelian randomization analysis with the European ancestry CKDGen data.

## Discussion

In this study, MR analysis demonstrated a causal linkage between NAFLD and eGFR. When the UK Biobank population was stratified according to the presence of the G allele of rs738409 or those with an increased genetic predisposition for NAFLD, which would lead to an increased prevalence of NAFLD in these individuals, showed lower eGFR. However, when the analysis was limited to those with a reduced possibility of developing NAFLD, the linkage was not identified, indicating that the genetic effect is actually through NAFLD. The significant association between genetic predisposition for NAFLD and lower eGFR was replicated in independent CKDGen GWAS meta-analysis summary data which has the largest scale to date.

There have been a number of reports suggesting that the presence of NAFLD is associated with an increased risk of CKD.^3^ However, as NAFLD itself primarily develops in individuals with metabolic disorders, which also cause kidney function impairment, the possibility that the findings are affected by reverse causation or residual confounding effects remains because of the inevitable limitation of observational studies. In this study, we performed an MR analysis that is known to be minimally affected by such issues and demonstrated the causal linkage between NAFLD and reduced eGFR. This is a large-scale finding that confirms a previously suggested association between the PNPLA3 variant rs738409 and reduced eGFR in two independent population-scale genetic datasets. In addition, with our efforts to attain the MR assumption, we demonstrated that the causal linkage would occur through NAFLD.

The study paid particular attention to the exclusion-restriction assumption of MR, as previous studies reported that rs738409 may directly affect kidney function independent of the presence of NAFLD.^6,8,9^ However, as the majority of studies included NAFLD diagnosed by imaging techniques as an adjustment variable, some undiagnosed or subclinical NAFLD cases might have been present. In addition, as such studies included only a few hundred individuals, the possibility of selection bias remained. As we stated, the presence of an independent effect of rs738409 on kidney function cannot explain the finding that the association was absent in those with a reduced risk of developing NAFLD. In addition, considering that the association strength was much weaker with kidney function than it was with NAFLD, it is reasonable to interpret that the association is mediated by NAFLD.^18^ In addition, that rs738409 is associated with reduced kidney function in within those with NAFLD may be because the genetic variant is associated with the severity of NAFLD,^5,29^ and severe NAFLD would have a larger effect on kidney function.

The causal linkage between liver cirrhosis and kidney function impairment has been suggested in past studies.^30^ A hepatic failure state has been recognized to cause “hepatorenal syndrome”, and such severe kidney failure due to advanced liver cirrhosis has been reported to be reversible by recovering liver function through transplantation. The suggested mechanisms include loss of effect volume by splanchnic vasodilation, increased inflammatory responses related or unrelated to bacterial translocation, and cirrhotic cardiomyopathy, which can be caused by cirrhosis. As NAFLD is a causal factor for liver cirrhosis in the advanced state, the causal effect of NAFLD on kidney function could be explained by this mechanism and such effect may be present even in the early stages of hepatic cirrhosis. The remaining question is the threshold severity of NAFLD that may significantly affect kidney function, which requires a future clinical study to be answered.

The clinical implication of this study is that, as NAFLD is a causal factor for reduced eGFR, kidney function should be monitored in NAFLD patients. Further, as CKD is an important risk factor for all-cause mortality and cardiovascular diseases,^2^ such a state should be promptly diagnosed, and early intervention to delay the progression of kidney function impairment should be considered in NAFLD patients experiencing eGFR loss. In addition, screening for NAFLD may be considered in those with CKD, as progression of NAFLD may accelerate kidney function decline. Individuals with common risk factors, such as obesity, hypertension, or diabetes, may be the particular targets of such screening strategies. Further, future clinical trials may ask whether management of NAFLD is beneficial for the risk of kidney function impairment development.

There are some limitations in this study. First, the currently identified magnitude of the effect size may differ in the clinical situation because of the nature of MR.^31^ In addition, as shared risk factors for metabolic comorbidities are common between NAFLD and CKD, the size of the observational association between NAFLD and CKD would be even larger than that reported in the current findings. Second, this study cannot directly prove that appropriate management of NAFLD would reduce the risk of kidney function impairment. Such clinical utility should be shown in a future clinical trial. Third, the included individuals were limited to individuals of European ancestry because the genetic distribution of SNPs may be different according to ethnic background, which would cause bias in the MR analysis. Thus, generalizability should be broadened in a future study investigating other ethnic populations. Last, the construction of a complete negative control was impossible, as NAFLD also occurs in those with a relatively healthy metabolic state. However, if we stringently define a subgroup with no risk or excluding phenotypical NAFLD, this would increase the possibility of collider bias.^32^

In conclusion, this MR analysis supports that NAFLD causally reduces eGFR. Clinicians should consider the causal linkage between NAFLD and kidney function impairment.

## Data Availability

The data for this study is available in the public domain.

## Acknowledgements

The study was based on the data provided by the UK Biobank consortium (application No. 53799). We thank the investigators of the previous studies who provided the summary statistics for the genetic instrument and the outcome of this study.

## Authors contributions

The corresponding author attests that all listed authors meet the authorship criteria and that no others meeting the criteria have been omitted. SP, HL, KSK, KWJ, and DKK contributed to the conception and design of the study. SL, YK, SC, YCK, SL, EKC, SSH, JPL, KWJ, CSL, YSK, and DKK provided statistical advice and interpreted the data. SP and KSK performed the main statistical analysis, assisted by SL and YK. HL, JPL, KWJ, CSL, YSK, and DKK provided advice regarding the data interpretation. YCK, SSH, HL, JPL, KWJ, CSL, and YSK provided material support during the study. All authors participated in drafting the manuscript. All authors reviewed the manuscript and approved the final version to be published.

## Data availability

The data for this study is available in the public domain described in the manuscript.

## List of abbreviations

NAFLD: non-alcoholic fatty liver diesase
CKD: chronic kidney disease
eGFR: estimated glomerular filtration rate
MR: Mendelian randomization
PGS: polygenic score

## References

1. Bellentani S. The epidemiology of non-alcoholic fatty liver disease. Liver Int. 2017;37 Suppl 1:81–84.

2. Global, regional, and national burden of chronic kidney disease, 1990-2017: a systematic analysis for the Global Burden of Disease Study 2017. Lancet. 2020;395:709–733.

3. Mantovani A, Zaza G, Byrne CD, Lonardo A, Zoppini G, Bonora E, et al. Nonalcoholic fatty liver disease increases risk of incident chronic kidney disease: A systematic review and meta-analysis. Metabolism. 2018;79:64–76.

4. Byrne CD, Targher G. NAFLD as a driver of chronic kidney disease. J Hepatol. 2020;72:785–801.

5. Anstee QM, Darlay R, Cockell S, Meroni M, Govaere O, Tiniakos D, et al. Genome-wide association study of non-alcoholic fatty liver and steatohepatitis in a histologically characterised cohort. J Hepatol. 2020;73:505–515.

6. Oniki K, Saruwatari J, Izuka T, Kajiwara A, Morita K, Sakata M, et al. Influence of the PNPLA3 rs738409 Polymorphism on Non-Alcoholic Fatty Liver Disease and Renal Function among Normal Weight Subjects. PLoS One. 2015;10:e0132640.

7. Musso G, Cassader M, Gambino R. PNPLA3 rs738409 and TM6SF2 rs58542926 gene variants affect renal disease and function in nonalcoholic fatty liver disease. Hepatology. 2015;62:658–659.

8. Mantovani A, Zusi C, Sani E, Colecchia A, Lippi G, Zaza GL, et al. Association between PNPLA3rs738409 polymorphism decreased kidney function in postmenopausal type 2 diabetic women with or without non-alcoholic fatty liver disease. Diabetes Metab. 2019;45:480–487.

9. Sun DQ, Zheng KI, Xu G, Ma HL, Zhang HY, Pan XY, et al. PNPLA3 rs738409 is associated with renal glomerular and tubular injury in NAFLD patients with persistently normal ALT levels. Liver Int. 2020;40:107–119.

10. Mantovani A, Taliento A, Zusi C, Baselli G, Prati D, Granata S, et al. PNPLA3 I148M gene variant and chronic kidney disease in type 2 diabetic patients with NAFLD: Clinical and experimental findings. Liver Int. 2020;40:1130–1141.

11. Davies NM, Holmes MV, Davey Smith G. Reading Mendelian randomisation studies: a guide, glossary, and checklist for clinicians. BMJ. 2018;362:k601.

12. Park S, Lee S, Kim Y, Lee Y, Kang MW, Kim K, et al. Short or Long Sleep Duration and CKD: A Mendelian Randomization Study. J Am Soc Nephrol. 2020;31:2937–2947.

13. Kennedy OJ, Pirastu N, Poole R, Fallowfield JA, Hayes PC, Grzeszkowiak EJ, et al. Coffee Consumption and Kidney Function: A Mendelian Randomization Study. Am J Kidney Dis. 2020;75:753–761.

14. Park S, Lee S, Kim Y, Lee Y, Kang MW, Kim K, et al. Causal effects of relative fat, protein, and carbohydrate intake on chronic kidney disease: a Mendelian randomization study. Am J Clin Nutr. 2021 [Epub ahead of print]

15. Park S, Lee S, Kim Y, Lee Y, Kang MW, Kim K, et al. Causal Effects of Positive Affect, Life Satisfaction, Depressive Symptoms, and Neuroticism on Kidney Function: A Mendelian Randomization Study. J Am Soc Nephrol. 2021 [Epub ahead of print]

16. Vujkovic M, Ramdas S, Lorenz KM, Schneider CV, Park J, Lee KM, et al. A genome-wide association study for nonalcoholic fatty liver disease identifies novel genetic loci and trait-relevant candidate genes in the Million Veteran Program. medRxiv. 2021:2020.2012.2026.20248491. [preprint, last accessed 2021-04-02]

17. Serper M, Vujkovic M, Kaplan DE, Carr RM, Lee KM, Shao Q, et al. Validating a non-invasive, ALT-based non-alcoholic fatty liver phenotype in the million veteran program. PLoS One. 2020;15:e0237430.

18. Hemani G, Tilling K, Davey Smith G. Orienting the causal relationship between imprecisely measured traits using GWAS summary data. PLoS Genet. 2017;13:e1007081.

19. Hemani G, Bowden J, Davey Smith G. Evaluating the potential role of pleiotropy in Mendelian randomization studies. Hum Mol Genet. 2018;27:R195–r208.

20. Fry A, Littlejohns TJ, Sudlow C, Doherty N, Adamska L, Sprosen T, et al. Comparison of Sociodemographic and Health-Related Characteristics of UK Biobank Participants With Those of the General Population. Am J Epidemiol. 2017;186:1026–1034.

21. Bycroft C, Freeman C, Petkova D, Band G, Elliott LT, Sharp K, et al. The UK Biobank resource with deep phenotyping and genomic data. Nature. 2018;562:203–209.

22. Lees JS, Welsh CE, Celis-Morales CA, Mackay D, Lewsey J, Gray SR, et al. Glomerular filtration rate by differing measures, albuminuria and prediction of cardiovascular disease, mortality and end-stage kidney disease. Nat Med. 2019;25:1753–1760.

23. Wuttke M, Li Y, Li M, Sieber KB, Feitosa MF, Gorski M, et al. A catalog of genetic loci associated with kidney function from analyses of a million individuals. Nat Genet. 2019;51:957–972.

24. Bowden J, Davey Smith G, Burgess S. Mendelian randomization with invalid instruments: effect estimation and bias detection through Egger regression. Int J Epidemiol. 2015;44:512–525.

25. Bowden J, Davey Smith G, Haycock PC, Burgess S. Consistent Estimation in Mendelian Randomization with Some Invalid Instruments Using a Weighted Median Estimator. Genet Epidemiol. 2016;40:304–314.

26. Burgess S, Foley CN, Allara E, Staley JR, Howson JMM. A robust and efficient method for Mendelian randomization with hundreds of genetic variants. Nat Commun. 2020;11:376.

27. Verbanck M, Chen CY, Neale B, Do R. Detection of widespread horizontal pleiotropy in causal relationships inferred from Mendelian randomization between complex traits and diseases. Nat Genet. 2018;50:693–698.

28. Hemani G, Zheng J, Elsworth B, Wade KH, Haberland V, Baird D, et al. The MR-Base platform supports systematic causal inference across the human phenome. Elife. 2018;7.

29. Parisinos CA, Wilman HR, Thomas EL, Kelly M, Nicholls RC, McGonigle J, et al. Genome-wide and Mendelian randomisation studies of liver MRI yield insights into the pathogenesis of steatohepatitis. J Hepatol. 2020;73:241–251.

30. Ginès P, Solà E, Angeli P, Wong F, Nadim MK, Kamath PS. Hepatorenal syndrome. Nat Rev Dis Primers. 2018;4:23.

31. Burgess S, Butterworth A, Malarstig A, Thompson SG. Use of Mendelian randomisation to assess potential benefit of clinical intervention. BMJ. 2012;345:e7325.

32. Taylor AE, Davies NM, Ware JJ, VanderWeele T, Smith GD, Munafò MR. Mendelian randomization in health research: using appropriate genetic variants and avoiding biased estimates. Econ Hum Biol. 2014;13:99–106.

