## Supplemental Table 1, Supplemental Table 2, Supplemental Method for "Causal effects from non-alcoholic fatty liver disease on kidney function: A Mendelian randomization study"

**Supplemental Methods.**

***Information collected in the UK Biobank data***

The information of the data from the UK Biobank consortium are available online (URL: https://www.ukbiobank.ac.uk/data-showcase/), and the information is identified by field IDs.

Baseline eGFR values in the UK Biobank were calculated from the information of cystatin C levels (Field ID 10720), along with information of ethnicity (Field ID 21000), calculated by the CKD-EPI equation. Other information was collected by as follows (the numbers of missing values in the clinical analysis dataset): age (Field ID 21003), sex (Field ID 31), smoking history (Field ID 20116), body mass index (Field ID 21001), waist circumference (Field ID 48), total cholesterol (Field ID 30690), HDL cholesterol (Field ID 30760), LDL cholesterol (Field ID 30780), and triglyceride levels (Field ID 30870). Self-reported history of treatment with hypertension medication or dyslipidemia medication (Field ID 6177 and 6153) was collected. A history of diabetes mellitus was collected by self-report, indicating a diabetes diagnosed by doctor (Field ID 2443). Systolic and diastolic BP was determined by the average of two automated measurements (Field ID 4080, and Field ID 4079), and those with a single missing measurement were considered to have missing information. Average total household income before tax (Field ID 738) which was graded was collected.

***Source of the genetic instrument of the group of variants and methods***

There has been a GWAS for histologically confirmed NAFLD, and the study reported 6 genetic loci that were strongly associated with NAFLD [1]. However, we identified that most of the loci were also strongly associated with obesity or metabolic syndrome-related traits; thus, the SNPs were not suitable for an MR study because of the issue of the horizontal pleiotropic pathway. Therefore, we first performed a single-variant MR analysis with rs738409, which has been repeatedly confirmed to be the SNP with the strongest association strength with NAFLD without low evidence of a pleiotropic effect. Selecting a relevant single variant as a genetic instrument has strength in MR, as the possibility of a pleiotropic effect is minimal, and such strength is important in our study, as kidney function and NAFLD are both largely affected by obesity-related traits.

To construct a genetic instrument for NAFLD that includes more relevant genetic variants, we needed a larger-scale GWAS identifying a larger number of SNPs with less evidence of a pleiotropic effect. Thus, we implemented the results from a preprint manuscript by Vojkovic M et al. (Posted January 02, 2021), which described a GWAS of clinically diagnosed NAFLD in a Million Veteran Program cohort [2]. The study implemented a clinically diagnosed NAFLD phenotype defined by (i) elevated ALT level >40 U/L for men and >30 U/L for women measured at at least two time points at least 6 months apart within a two-year window at any point prior to enrollment and (ii) exclusion of other causes of liver disease (e.g., presence of chronic viral hepatitis B or C, chronic liver diseases or systemic conditions, and/or alcohol use disorder). The control group in the study had a normal ALT level (≤30 U/L for men, ≤20 U/L for women) and no other causes of liver disease or alcohol use disorder or related conditions. The clinical diagnosis of NAFLD in the Million Veteran Program cohort has been validated in a previous study [3].

The study also reported the results from phenome-wide association studies in the UK Biobank with the reported genome-wide significance SNPs. The study performed the analysis with electronic health record-derived ICD diagnostic codes from UK Biobank participants with white British ancestry. The study screened 1403 traits and categorized them into relevant systems. We implemented the information and disregarded the SNPs that showed strong ($P < 0.001$) associations with the circulatory system or endocrine/metabolism-related traits, as the systems were considered to have possible pleiotropic effects in our MR analysis.

**Supplemental Table 1.** Genetic instrument for NAFLD reported in the genome-wide association study with the European ancestyr individuals of Million Veteran Program cohort.

| SNP | Chr | Position | effect allele | other allele | effect allele frequency | beta | standard error | P value | Remarks |
|---|---|---|---|---|---|---|---|---|---|
| rs1497406 | 1 | 16505320 | A | G | 0.426 | -0.042 | 0.007 | 1.46E-08 | |
| rs79598313 | 1 | 27284913 | T | C | 0.023 | 0.180 | 0.025 | 3.37E-13 | Included in the individual-level in the UK Biobank but not in the summary-level MR because of non-overlapping. |
| rs74816838 | 1 | 161643560 | T | C | 0.112 | 0.087 | 0.013 | 1.09E-11 | Included in the individual-level in the UK Biobank but not in the summary-level MR because of non-overlapping. |
| rs1337101 | 1 | 219726100 | T | G | 0.300 | -0.051 | 0.008 | 1.95E-10 | Disregarded: strong association with a confounder in "Circulatory system" or "Endocrine/metabolism" category. |
| rs2642438 | 1 | 220970028 | A | G | 0.295 | -0.079 | 0.008 | 8.04E-23 | |
| rs848559 | 2 | 36694497 | T | A | 0.151 | -0.061 | 0.011 | 7.59E-09 | |
| rs10195619 | 2 | 112765562 | T | C | 0.377 | -0.049 | 0.008 | 8.78E-11 | |
| rs13409360 | 2 | 113838102 | A | G | 0.397 | -0.060 | 0.008 | 1.95E-15 | |
| rs6717858 | 2 | 165539661 | C | T | 0.400 | -0.051 | 0.008 | 2.04E-11 | |
| rs73024760 | 2 | 169885122 | T | C | 0.038 | 0.107 | 0.020 | 4.64E-08 | Disregarded: strong association with a confounder in "Circulatory system" or "Endocrine/metabolism" category. |
| rs10201587 | 2 | 202202791 | G | A | 0.499 | -0.045 | 0.007 | 4.11E-10 | |
| rs2138157 | 2 | 227103717 | A | C | 0.353 | -0.064 | 0.008 | 3.49E-17 | |
| rs7604422 | 2 | 233509761 | C | A | 0.401 | -0.053 | 0.008 | 1.24E-12 | |
| rs4684847 | 3 | 12386337 | T | C | 0.123 | -0.072 | 0.011 | 1.11E-10 | |
| rs9867368 | 3 | 136147771 | A | G | 0.198 | -0.078 | 0.009 | 1.61E-17 | |
| rs9833411 | 3 | 142640398 | A | T | 0.357 | -0.048 | 0.008 | 1.08E-09 | |
| rs1594320 | 3 | 149114757 | G | C | 0.296 | 0.053 | 0.008 | 5.25E-11 | |
| rs12486792 | 3 | 170677123 | G | C | 0.136 | 0.060 | 0.011 | 2.95E-08 | |
| rs574044675 | 3 | 172274232 | C | A | 0.024 | -0.251 | 0.027 | 5.07E-20 | Included in the individual-level in the UK Biobank but not in the summary-level MR because of non-overlapping. |
| rs12500824 | 4 | 77416627 | A | G | 0.350 | 0.046 | 0.008 | 1.37E-09 | Disregarded: reverse direction effect and strong association with eGFR (P = 2.61E-61) and association with a confounder. |
| rs71633358 | 4 | 88183817 | C | T | 0.281 | -0.090 | 0.009 | 1.89E-26 | Included in the individual-level in the UK Biobank but not in the summary-level MR because of non-overlapping. |
| rs4148824 | 7 | 87075362 | G | A | 0.185 | -0.057 | 0.010 | 2.62E-09 | |
| rs4841133 | 8 | 9183664 | A | G | 0.088 | 0.130 | 0.013 | 1.02E-24 | |
| rs4484649 | 8 | 10571491 | C | A | 0.413 | 0.045 | 0.008 | 2.16E-09 | Disregarded because of its' strong association with a confounder in "Circulatory system" or "Endocrine/metabolism" category. |
| rs4734654 | 8 | 103669991 | G | A | 0.362 | -0.052 | 0.008 | 1.48E-11 | |
| rs2954038 | 8 | 126507389 | C | A | 0.306 | 0.139 | 0.008 | 2.12E-70 | |
| rs147998249 | 8 | 145732180 | C | G | 0.003 | -2.019 | 0.123 | 7.52E-61 | Included in the individual-level in the UK Biobank but not in the summary-level MR because of non-overlapping. |
| rs7041363 | 9 | 117146043 | G | C | 0.491 | -0.135 | 0.008 | 1.12E-71 | |
| rs10883451 | 10 | 101924418 | C | T | 0.477 | -0.161 | 0.007 | 3.60E-106 | |
| rs17780834 | 10 | 104091096 | T | A | 0.061 | 0.086 | 0.015 | 1.65E-08 | |
| rs2792751 | 10 | 113940329 | T | C | 0.293 | 0.072 | 0.008 | 1.28E-19 | |
| rs11601507 | 11 | 5701074 | A | C | 0.073 | 0.088 | 0.014 | 2.16E-10 | Disregarded: strong association with a confounder in "Circulatory system" or "Endocrine/metabolism" category. |
| rs174535 | 11 | 61551356 | C | T | 0.337 | -0.061 | 0.008 | 3.11E-15 | Disregarded: strong association with a confounder in "Circulatory system" or "Endocrine/metabolism" category. |
| rs7117339 | 11 | 93870338 | T | C | 0.120 | -0.130 | 0.011 | 2.26E-30 | |
| rs4919741 | 12 | 53272920 | A | G | 0.344 | -0.057 | 0.008 | 1.72E-13 | |
| rs1169292 | 12 | 121426478 | T | C | 0.321 | 0.054 | 0.008 | 4.45E-12 | |
| rs148015593 | 12 | 122523668 | G | T | 0.477 | -0.042 | 0.007 | 8.69E-09 | |
| rs11621792 | 14 | 24871926 | T | C | 0.453 | 0.042 | 0.007 | 1.11E-08 | Disregarded: strong association with a confounder in "Circulatory system" or "Endocrine/metabolism" category. |
| rs28929474 | 14 | 94844947 | T | C | 0.018 | 0.481 | 0.028 | 1.03E-65 | Disregarded: strong association with a confounder in "Circulatory system" or "Endocrine/metabolism" category. |
| rs168144 | 15 | 60914262 | C | T | 0.433 | -0.052 | 0.007 | 3.45E-12 | |
| rs55868793 | 15 | 73956856 | G | T | 0.408 | 0.059 | 0.008 | 8.92E-15 | |
| rs72754571 | 15 | 90350888 | A | C | 0.081 | -0.083 | 0.014 | 2.47E-09 | |
| rs112128680 | 16 | 72054052 | A | G | 0.149 | 0.061 | 0.010 | 3.70E-09 | |
| rs4782568 | 16 | 83980529 | G | C | 0.438 | -0.064 | 0.008 | 9.17E-18 | |
| rs1801689 | 17 | 64210580 | C | A | 0.032 | 0.176 | 0.021 | 1.39E-17 | Disregarded: strong association with a confounder in "Circulatory system" or "Endocrine/metabolism" category. |
| rs4940689 | 18 | 56089116 | A | G | 0.205 | 0.054 | 0.010 | 1.21E-08 | |
| rs3810367 | 19 | 4342847 | G | T | 0.376 | 0.045 | 0.008 | 3.51E-09 | Disregarded: strong association with a confounder in "Circulatory system" or "Endocrine/metabolism" category. |
| rs58542926 | 19 | 19379549 | T | C | 0.075 | 0.222 | 0.014 | 2.13E-59 | Disregarded: strong association with a confounder in "Circulatory system" or "Endocrine/metabolism" category. |
| rs7599 | 19 | 36038390 | A | G | 0.382 | 0.049 | 0.008 | 5.19E-11 | |
| rs429358 | 19 | 45411941 | C | T | 0.139 | -0.094 | 0.011 | 4.72E-19 | |
| rs2377957 | 20 | 32554473 | A | G | 0.329 | -0.053 | 0.008 | 1.53E-11 | |
| rs2207132 | 20 | 39142516 | A | G | 0.029 | 0.189 | 0.029 | 7.96E-11 | Disregarded: strong association with a confounder in "Circulatory system" or "Endocrine/metabolism" category. |
| rs1547014 | 22 | 29100711 | T | C | 0.302 | -0.064 | 0.008 | 5.31E-16 | Disregarded: strong association with a confounder in "Circulatory system" or "Endocrine/metabolism" category. |
| rs132665 | 22 | 36564170 | G | A | 0.154 | -0.069 | 0.010 | 1.84E-11 | Disregarded: strong association with a confounder in "Circulatory system" or "Endocrine/metabolism" category. |
| rs738409 | 22 | 44324727 | G | C | 0.228 | 0.269 | 0.0086 | 2.84E-213 | |

The genetic instrument was developed from a previous large-scale GWAS for clinical NAFLD (Vujkovic M, et al. . medRxiv. 2021:2020.2012.2026.20248491. last accessed 2021-04-02).

**Supplemental Table 2.** The SNPs that were disregarded because of strong association with possible confounders in "Circulatory system" or "endocrine/metabolism" category identified by phenome-wide association study in the UK Biobank

| SNP | Category | Phenotype | P value |
|---|---|---|---|
| rs11601507 | Circulatory System | Ischemic Heart Disease | 1.10E-05 |
| rs11601507 | Circulatory System | Angina pectoris | 5.90E-05 |
| rs11601507 | Circulatory System | Coronary atherosclerosis | 1.70E-04 |
| rs12500824 | Circulatory System | Myocardial infarction | 5.50E-05 |
| rs12500824 | Circulatory System | Essential hypertension | 8.80E-05 |
| rs12500824 | Circulatory System | Hypertension | 1.00E-04 |
| rs12500824 | Circulatory System | Coronary atherosclerosis | 4.30E-04 |
| rs132665 | Circulatory System | Phlebitis and thrombophlebitis of lower extremities | 1.50E-05 |
| rs132665 | Circulatory System | Phlebitis and thrombophlebitis | 1.10E-04 |
| rs132665 | Circulatory System | Nonspecific abnormal findings on radiological and other examination of other intrathoracic organs (echocardiogram, etc) | 1.70E-04 |
| rs132665 | Circulatory System | Essential hypertension | 2.90E-04 |
| rs132665 | Circulatory System | Hypertension | 3.40E-04 |
| rs1337101 | Circulatory System | Varicose veins of lower extremity | 8.70E-06 |
| rs1337101 | Circulatory System | Varicose veins | 8.50E-05 |
| rs1547014 | Circulatory System | Hypertension | 7.50E-04 |
| rs174535 | Circulatory System | Abnormal heart sounds | 6.30E-04 |
| rs1801689 | Circulatory System | Congestive heart failure; nonhypertensive | 9.50E-05 |
| rs1801689 | Circulatory System | Congestive heart failure (CHF) NOS | 2.70E-04 |
| rs1801689 | Circulatory System | Cardiac dysrhythmias | 5.00E-04 |
| rs2207132 | Circulatory System | Coronary atherosclerosis | 1.00E-04 |
| rs2207132 | Circulatory System | Angina pectoris | 5.40E-04 |
| rs2207132 | Circulatory System | Ischemic Heart Disease | 6.70E-04 |
| rs28929474 | Circulatory System | Coronary atherosclerosis | 3.00E-06 |
| rs28929474 | Circulatory System | Myocardial infarction | 8.10E-06 |
| rs28929474 | Circulatory System | Angina pectoris | 4.80E-05 |
| rs28929474 | Circulatory System | Ischemic Heart Disease | 7.90E-05 |
| rs28929474 | Circulatory System | Other chronic ischemic heart disease, unspecified | 2.60E-04 |
| rs28929474 | Circulatory System | Other aneurysm | 8.50E-04 |
| rs3810367 | Circulatory System | Bundle branch block | 9.00E-05 |
| rs3810367 | Circulatory System | Left bundle branch block | 4.70E-04 |
| rs3810367 | Circulatory System | Cardiac conduction disorders | 8.20E-04 |
| rs4484649 | Circulatory System | Hypertension | 4.50E-15 |
| rs4484649 | Circulatory System | Essential hypertension | 6.20E-15 |
| rs4484649 | Circulatory System | Orthostatic hypotension | 7.30E-04 |
| rs79598313 | Circulatory System | Essential hypertension | 5.80E-05 |
| rs79598313 | Circulatory System | Hypertension | 7.90E-05 |
| rs79598313 | Circulatory System | Ischemic Heart Disease | 3.60E-04 |
| rs79598313 | Circulatory System | Angina pectoris | 9.10E-04 |
| rs11601507 | Endocrine/Metabolism | Hypercholesterolemia | 4.50E-06 |
| rs11601507 | Endocrine/Metabolism | Hyperlipidemia | 7.90E-06 |
| rs11601507 | Endocrine/Metabolism | Disorders of lipoid metabolism | 9.20E-06 |
| rs11621792 | Endocrine/Metabolism | Hyperlipidemia | 2.10E-04 |
| rs11621792 | Endocrine/Metabolism | Disorders of lipoid metabolism | 2.30E-04 |
| rs11621792 | Endocrine/Metabolism | Hypercholesterolemia | 5.00E-04 |
| rs12500824 | Endocrine/Metabolism | Hyperaldosteronism | 5.10E-04 |
| rs1337101 | Endocrine/Metabolism | Diabetes mellitus | 2.70E-06 |
| rs1337101 | Endocrine/Metabolism | Type 2 diabetes | 5.50E-06 |
| rs174535 | Endocrine/Metabolism | Hypothyroidism | 9.40E-06 |
| rs174535 | Endocrine/Metabolism | Hypothyroidism NOS | 1.30E-05 |
| rs1801689 | Endocrine/Metabolism | Hypercholesterolemia | 1.60E-05 |
| rs1801689 | Endocrine/Metabolism | Disorders of lipoid metabolism | 1.60E-04 |
| rs1801689 | Endocrine/Metabolism | Hyperlipidemia | 1.70E-04 |
| rs2207132 | Endocrine/Metabolism | Hyperlipidemia | 1.80E-11 |
| rs2207132 | Endocrine/Metabolism | Disorders of lipoid metabolism | 2.00E-11 |
| rs2207132 | Endocrine/Metabolism | Hypercholesterolemia | 1.00E-10 |
| rs2207132 | Endocrine/Metabolism | Mixed hyperlipidemia | 5.60E-05 |
| rs28929474 | Endocrine/Metabolism | Other disorders of metabolism | 1.10E-13 |
| rs28929474 | Endocrine/Metabolism | Hyperlipidemia | 1.90E-04 |
| rs28929474 | Endocrine/Metabolism | Disorders of lipoid metabolism | 2.20E-04 |
| rs4484649 | Endocrine/Metabolism | Obesity | 3.70E-05 |
| rs4484649 | Endocrine/Metabolism | Overweight, obesity and other hyperalimentation | 7.50E-05 |
| rs4484649 | Endocrine/Metabolism | Type 2 diabetes | 8.90E-05 |
| rs4484649 | Endocrine/Metabolism | Diabetes mellitus | 7.30E-04 |
| rs58542926 | Endocrine/Metabolism | Hypercholesterolemia | 1.00E-15 |
| rs58542926 | Endocrine/Metabolism | Disorders of lipoid metabolism | 6.10E-15 |
| rs58542926 | Endocrine/Metabolism | Hyperlipidemia | 1.20E-14 |
| rs58542926 | Endocrine/Metabolism | Diabetes mellitus | 1.10E-06 |
| rs58542926 | Endocrine/Metabolism | Type 2 diabetes | 1.40E-06 |
| rs58542926 | Endocrine/Metabolism | Diabetes insipidus | 2.50E-04 |
| rs73024760 | Endocrine/Metabolism | Polycystic ovaries | 5.20E-04 |
| rs79598313 | Endocrine/Metabolism | Disorders of lipoid metabolism | 2.10E-07 |
| rs79598313 | Endocrine/Metabolism | Hyperlipidemia | 2.20E-07 |
| rs79598313 | Endocrine/Metabolism | Hypercholesterolemia | 2.70E-07 |
